# Transmission of SARS-CoV-2 within households: a prospective cohort study in the Netherlands and Belgium – Interim results

**DOI:** 10.1101/2021.04.23.21255846

**Authors:** JDM Verberk, MLA de Hoog, I Westerhof, S Van Goethem, C Lammens, M Ieven, E de Bruin, J Bielicki, S Coenen, J van Beek, M Bonten, H Goossens, PCJL Bruijning-Verhagen

## Abstract

**Background:** Household transmission studies are useful to obtain granular data on SARS-CoV-2 transmission dynamics and to gain insight into the main determinants. In this interim report we investigated secondary attack rates (SAR) by household and subject characteristics in the Netherlands and Belgium.

**Methods:** Households with a real-time reverse transcription polymerase chain reaction (RT-PCR) confirmed SARS-CoV-2 index case were enrolled <48 hours following report of the positive test result. Daily symptom follow-up, standardized nose-throat sampling at enrollment and at new-onset acute respiratory illness (ARI) and paired dried blood spots (DBS) were collected from each participant. Children 0-2 years of age were additionally requested to collect a stool sample 7 days after enrollment and at new-onset of ARI. Swabs and stool samples were tested by RT-PCR for virus detection and DBS by multiplex protein microarray for detection of SARS-CoV-2 antibodies. The SAR was calculated 1) per-household as the proportion of households with ≥1 secondary SARS-CoV-2 case and 2) per-person as the probability of infection in household members at risk. We explored differences in SARs by household and subject characteristics.

**Results:** This analysis includes 117 households that completed follow-up between April-December 2020. Among 382 subjects, 74 secondary infections were detected, of which 13 (17.6%) were asymptomatic and 20 (27.0%) infections were detected by seroconversion only. Of cases detected by RT-PCR, 50 (67.6%) were found at enrollment. The household SAR was 44.4% (95%-CI: 35.4-53.9%) and was higher for index cases meeting the ARI case definition (52.3%; 95%-CI 41.4-62.9%) compared to mildly symptomatic (22.2%; 95%-CI: 9.4-42.7%) and asymptomatic index cases (0.0%; 95%-CI: 0.0-80.2%). The per-person SAR was 27.9% (95%-CI: 22.7-33.8%). Transmission was lowest from child to parent (9.1%; 95%-CI: 2.4-25.5%) and highest from parent to child (28.1%; 95%-CI: 19.7-38.4%) and in children 6-12 years (34.2%; 95%-CI: 20.1-51.4%). Among 141 subjects with RT-PCR confirmed SARS-CoV-2 infections, seroconversion was detected in 111 (78.7%).

**Conclusion:** We found a high household SAR, with the large majority of transmissions detected early after identification of the index case. Our findings confirm differential SAR by symptom status of the index. In almost a quarter of RT-PCR positive cases, no antibodies were detected. Other factors influencing transmission will be further explored as more data accumulate.

## INTRODUCTION

Despite the widespread implementation of severe acute respiratory syndrome coronavirus 2 (SARS-CoV-2) infection control guidelines to prevent transmission within households, households remain the most important sites for disease transmission and are therefore considered an important factor in sustaining the pandemic.[1, 2] Madewell et al. [3] performed a meta-analysis of household studies conducted in the first six months of the pandemic, mostly Chinese studies, and found an average household secondary attack rate (SAR) of 16.6% (95%CI: 14.0-19.3%), but percentages varied widely across studies. The majority of the household studies published so far, however, are retrospective, rely on symptomatic cases only, or only use real-time reverse transcription polymerase chain reaction (RT-PCR) to identify cases. As SARS-CoV-2 infection can present with mild symptoms or completely asymptomatically, in particular in younger individuals, this methodology may underestimate the true within household transmission. A Dutch household study among 55 households with children found a much higher household SAR of 43% (95%-CI: 33-53%), presumably owing to a dense sampling protocol testing both symptomatic and asymptomatic individuals with regular RT-PCR and serology testing.[4] Household transmission may also depend on socio-cultural factors and living conditions, therefore results may not be generalizable between settings or regions. So far, limited data are available from households in Western Europe.

In April 2020, we initiated a prospective study in the Netherlands (NL), Belgium (BE) and Switzerland (CH), to closely monitor household members of confirmed SARS-CoV-2 index patients with varying disease severity and demographic characteristics in order to estimate SAR in Western European setting and to quantify key household transmission characteristics for both symptomatic and asymptomatic SARS-CoV-2 infections. In this report, we include interim results based on data collected in Belgium (BE) and the Netherlands (NL) from April 20, 2020 to December 2, 2021.

## METHODS

### Study design and data collection

Within this prospective cohort study, households consisting of at least two members were recruited by the University Medical Centre Utrecht (NL) and the University of Antwerp (BE), either via healthcare worker screening programs for SARS-CoV-2, drive-through testing sites, general practitioner visits or pre-operative screening programs. Households were eligible following a first laboratory-confirmed positive SARS-CoV-2 test result in a household member (index case) and enrolled within 48 hours following test result. Index cases could be either symptomatic or asymptomatic as long as they received a positive RT-PCR result. Medical Ethical Committee Utrecht (NL) and Ethical Committee of University Hospital Antwerp (BE) provided review and ethical approval of the study (Reference number 20-185/D and 20/14/177 respectively). Written informed consent was obtained from all participating household members or their legal guardians.

Following enrollment each household member, including the index case, was instructed to take a nose-throat swab by self-sampling at home and a dried blood spot (DBS) by self-finger-prick. A stool sample on day seven was included for children aged 0-2 years. Self-sampling was supported by instruction videos and leaflets delivered with the sampling material. A telephone helpdesk was available 6 days a week during working hours. A baseline questionnaire was completed on household member characteristics and living conditions. Daily follow-up included a digital diary for each participating household member detailing respiratory and systemic symptoms. If reported symptoms met the case definition for acute respiratory illness (ARI), an additional nose-throat swab was requested from the symptomatic person (for symptomatic children 0-2 years a stool sample was also included seven days post-symptom onset). Follow-up ended after 21 days or 21 days following the last ARI onset in a household member. Ten days later a second DBS was collected from all household members. All data were collected by means of a study App (COVapp), which is a custom-made application compatible with Apple and Android systems, developed by the UMCU. All data entered in the App were stored in an online secured database.

### Laboratory analyses

Nose-throat swabs and stool samples were tested separately for the presence of SARS-CoV-2 by RT-PCR as described in detail elsewhere.[5, 6] Specimens with a cycle threshold (Ct) value less than or equal to 40 were defined as positive. A Ct-value that exceeds 40 was defined as a negative test.

DBS specimens were tested in a final dilution of 1:40 by multiplex protein microarray for IgG antibodies targeting recombinant SARS-CoV-2 spike (S) ectodomain and S1 domain subunit antigens expressed in HEK293 cells as described elsewhere.[7-9] The S1 antigen signal exceeding 13,000 relative fluorescence units (RFU) and a S ectodomain exceeding 2,000 RFU were considered positive. See Supplement for more details.

### Case definitions

An ARI episode based on daily symptom reports was defined as onset of fever <u>OR</u> two consecutive days with 1) at least two respiratory symptoms (cough, sore throat, cold, dyspnea) <u>OR</u> 2) one respiratory symptom combined with at least one systemic symptom (headache, muscle ache, cold shivers or fatigue). Any subject reporting respiratory or systemic symptoms, but not meeting the ARI case-definition was defined as mildly symptomatic.

SARS-CoV-2 infection was defined as 1) a positive RT-PCR sample (nose-throat or fecal); or 2) negative serology at enrollement and positive at end of follow-up (seroconversion). Asymptomatic SARS-CoV-2 infections included subjects without reported symptoms during follow-up but with either a positive RT-PCR result at enrollment and/or seroconversion. Similarly, symptomatic SARS-CoV-2 infections could be detected based on positive RT-PCR and/or seroconversion.

A secondary case was defined as any SARS-CoV-2 infection in a household member not being the index case, detected during follow-up.

### Statistical analysis

Baseline characteristics were described for households with and without secondary transmission and differences between the two groups were tested by t-test (continuous variables) or chi-square (categorical variables). First, we calculated the household SAR by dividing the number of households with at least one secondary case by the total number of participating households. Second, the per-person SAR was calculated by dividing the number of secondary SARS-CoV-2 cases by the number of household members at risk.[10] In addition, we explored household and individual characteristics of index cases and household members associated with SARS-CoV-2 transmission. In a sub-analysis we calculated the household and per-person SAR for symptomatic household members with positive RT-PCR results only, reflecting a restrictive testing policy which is mostly used in retrospective studies. Last, seroconversion rate was calculated for RT-PCR positive subjects with ARI, mild or no symptoms (index and secondary cases). Differences between the three groups were tested by chi-square.

In a sensitivity analysis all household members with a positive RT-PCR at enrollment were considered a co-primary index case. The household SAR was re-calculated excluding the households with only co-primary cases and per-person SAR was re-calculated excluding the co-primary cases from the household members at risk.

Household members were excluded from the analysis when their SARS-CoV-2 status could not be defined because no test results were available to determine absence or presence of SARS-CoV-2 infection (unknown secondary case status).

## RESULTS

In this ongoing study a total of 272 households are enrolled of which 155 households are still in follow-up or have samples awaiting lab analysis. Five household members were excluded because their secondary case status could not be defined. For this analysis, data were available for 117 households and 382 subjects (117 index cases and 265 household members; Figure 1). Supplement Table 1 provides an overview of the adherence to the sampling protocol and the number of missing serology results due to insufficient sampling. The majority of the 117 households (65%) was enrolled during the second epidemic wave (September-December 2020). Twenty households were enrolled in Belgium and 97 in the Netherlands. The median duration of follow-up per household was 35 days (IQR: 31-40 days).

**Figure 1:**
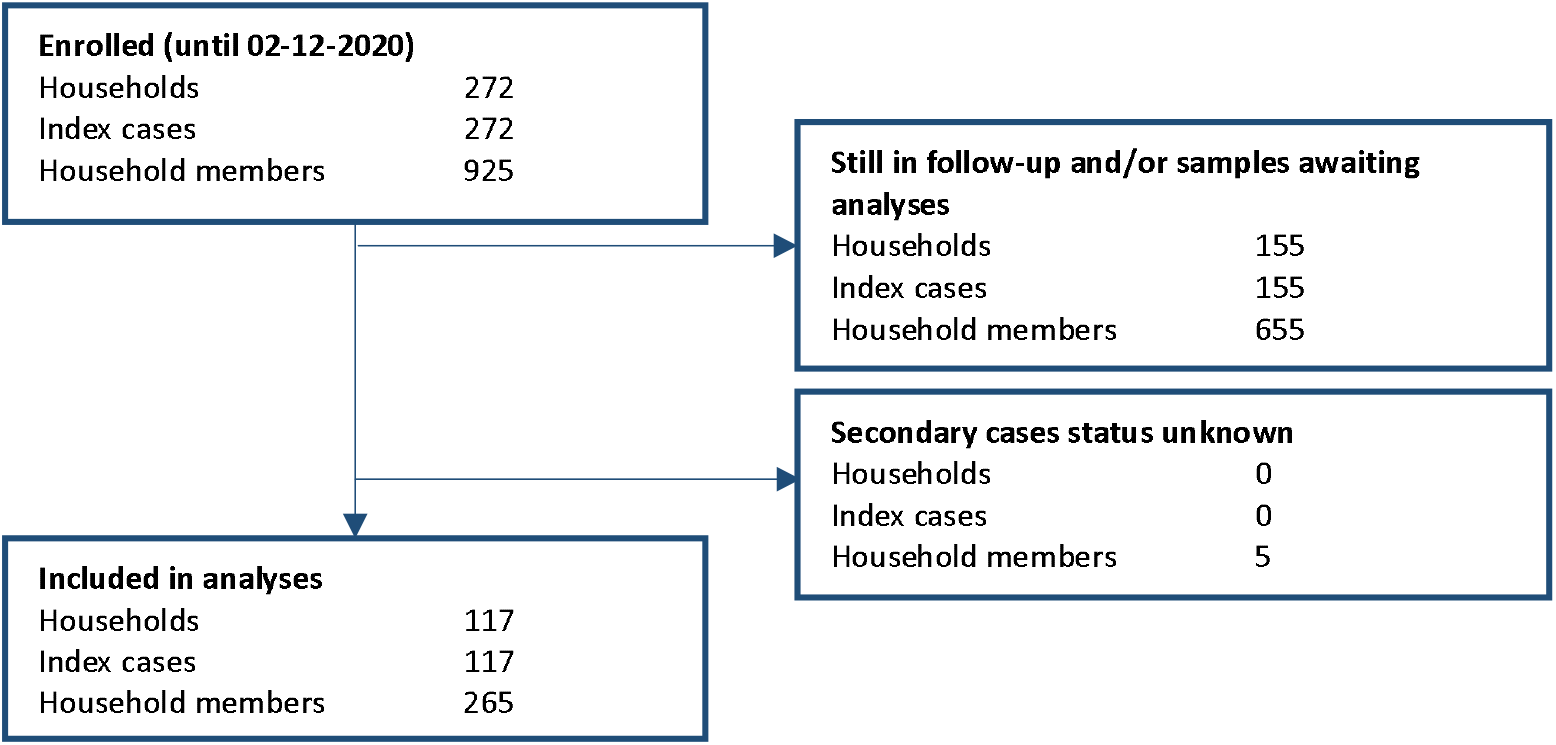
Flow diagram of the study population

Table 1 shows the characteristics of the 117 households and index cases. The median household size was 3 persons (IQR: 2-4). The median age of the index case was 39.0 years (IQR: 26.5-51.0 years); eight (6.9%) index cases were <18 years. Index cases were predominantly female (n=73; 62.4%). The majority of index cases met the ARI case definition (n=88, 75.2%) and only two (1.7%) were asymptomatic. The median time between symptom onset and positive test result of index cases was 3 days (IQR: 2-5 days) and study enrollment started after a median of 5 days (IQR: 3-6 days) after symptom onset.

**Table 1:**
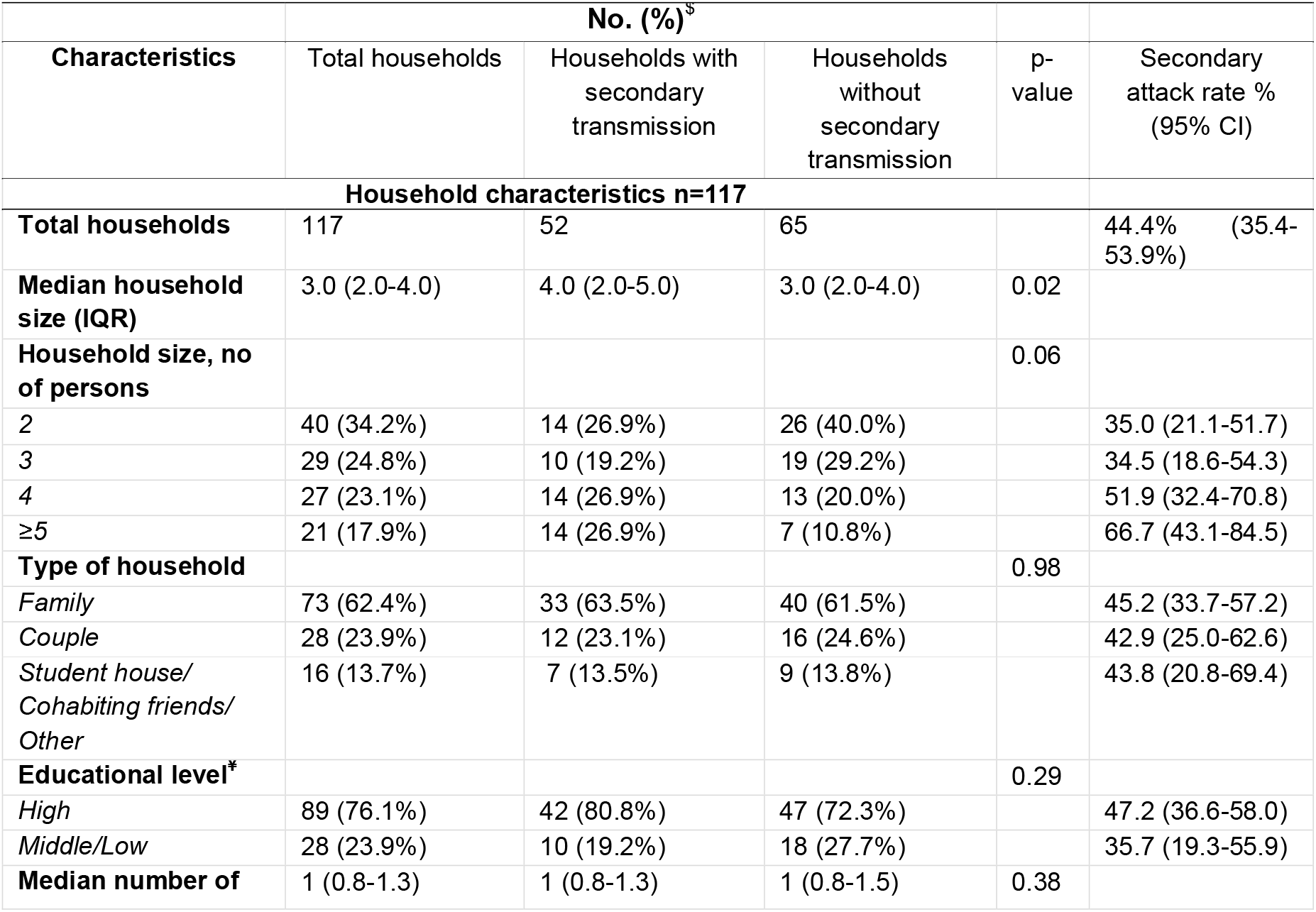
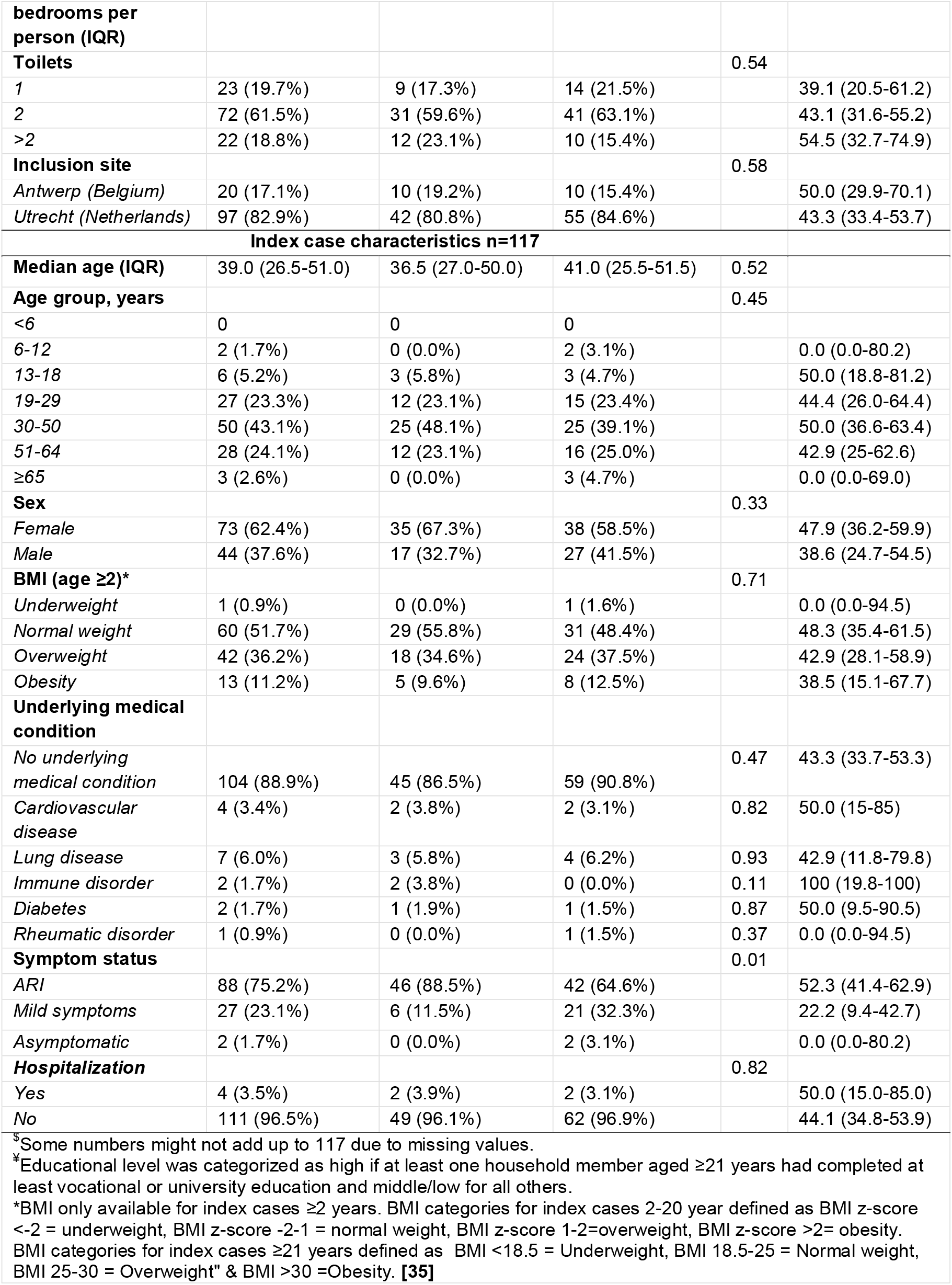
Characteristics of households and index cases (117 households)

Secondary transmission was detected in 52 out of 117 households (household SAR: 44.4%; 95%-CI: 35.4-53.9%). The household SAR was significantly higher (p=0.01) when the index had ARI (52.3%; 95%-CI: 41.4-62.9%) compared to mildly symptomatic (22.2%; 95%-CI: 9.4-42.7%) or completely asymptomatic index cases (0.0%; 95%-CI: 0.0-80.2%). The median household size was larger in households with secondary transmission compared to households without secondary transmission (median 4.0; IQR: 2.0-5.0 and 3.0; IQR 2.0-4.0 respectively; p=0.02). The detected household SAR would decline to 35.0% (95%-CI: 26.6-44.5%) when using a more restrictive testing policy of RT-PCR testing for symptomatic cases only, as used in most retrospective studies (Table2).

**Table 2:** Estimates of within-household transmission

| Parameter | % (95%CI) | n/N |
| --- | --- | --- |
| Household SAR† | 44.4 (35.4-53.9) | 52/117 |
| Household SAR symptomatic secondary cases with positive RT-PCR^ | 35.0 (26.6-44.5) | 41/117 |
| Household SAR asymptomatic secondary cases\$ | 9.4 (5.0-16.6) | 11/117 |
| Per-person SAR‡ | 27.9 (22.7-33.8) | 74/265 |
| Per-person SAR symptomatic secondary cases with positive RT-PCR# | 16.6 (12.4-21.8) | 44/265 |
| Per-person SAR asymptomatic secondary cases ¶ | 4.9 (2.8-8.4) | 13/265 |
| Proportion asymptomatic secondary cases§ | 17.6 (10.0-28.5) | 13/74 |

†Number of households with at least one secondary case divided by the number of households at risk. ^Including only symptomatic and RT-PCR positive secondary cases. \$Including only asymptomatic cases. ‡Number of household secondary cases divided by the number of household members at risk. # Including only symptomatic and RT-PCR positive secondary cases. ¶ Including only asymptomatic secondary cases. §Number asymptomatic divided by the total number of secondary cases.
†Number of households with at least one secondary case divided by the number of households at risk. ^Including only symptomatic and RT-PCR positive secondary cases. \$Including only asymptomatic cases. ‡Number of household secondary cases divided by the number of household members at risk.

Table 3 shows the characteristics of 265 household members. The median age of the household members was 24 years (IQR: 14.0-47.5 years); 96 (36.2%) household members were <18 years. Among all 265 household members, 10 (3.8%) showed antibodies against SARS-CoV-2 at enrollment of which in 4/10 (40%) subjects it was not accompanied with a positive RT-PCR, suggesting that they have been infected earlier.

**Table 3:**
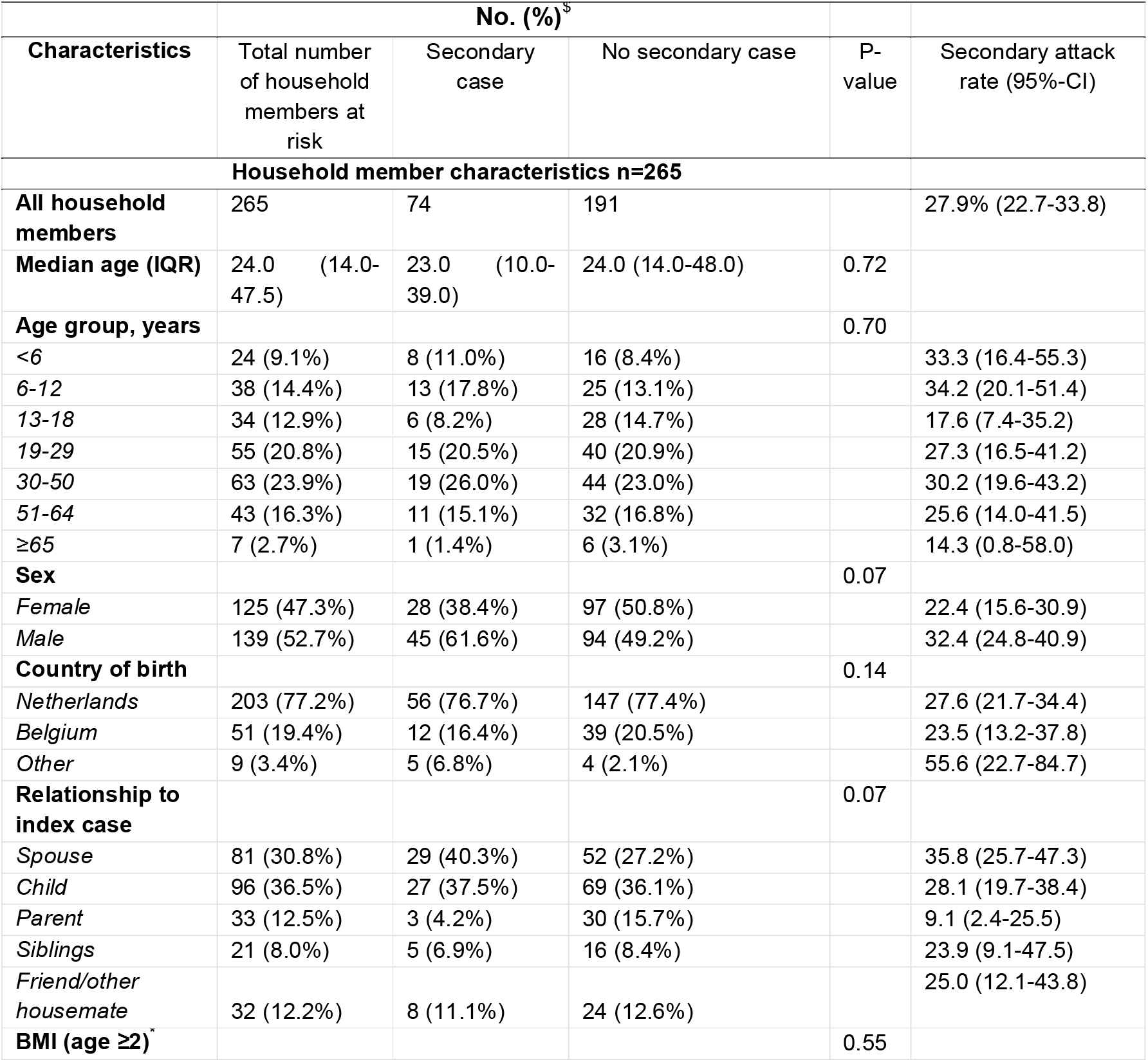
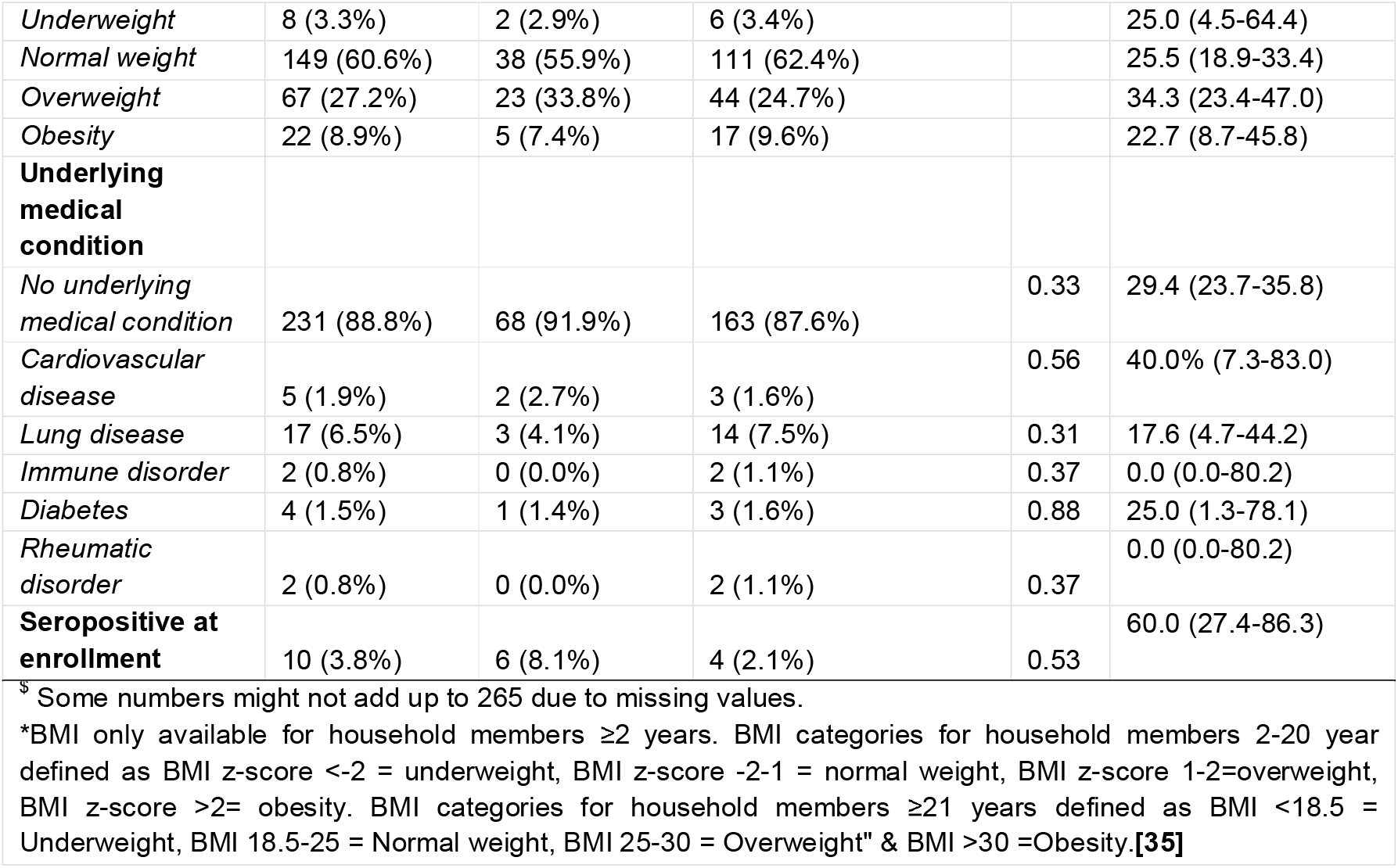
Characteristics of household members, n=265

Secondary transmission was detected in 74 of the 265 household members (per-person SAR: 27.9%; 95%-CI: 22.7-33.8%). The per-person SAR varied by the household member age-group between 14.3% (95%-CI: 0.8-58.0%; >65 years) and 34.2% (95%-CI:20.1-51.4%; 6-12 years). It was lowest from child index to parent (9.1%; 95%-CI: 2.4-25.5%) and highest from partner to spouse (35.8%; 95%-CI: 25.7-47.3%).

Table 4 shows the episode characteristics of the 74 household members with secondary transmission. Fifty (67.6%) secondary cases were already detected by RT-PCR at enrollment. Twenty (27.0%) infections were detected by seroconversion only. In three (4.1%) subjects, symptoms had already started in the two weeks before enrollment, indicating the direction of transmission may have been in opposite direction. An additional 45 subjects (60.8%) reported respiratory symptoms at enrollment. Overall, 42 (56.8%) secondary cases had ARI, 19 (25.7%) were mildly symptomatic and 13 (17.6%) were asymptomatic. A more restrictive testing policy of RT-PCR testing for symptomatic cases only would have yielded a per-person SAR of 16.6% (95%-CI: 12.4-21.8%; Table 2).

**Table 4:** Episode characteristics of the 74 secondary cases.

| No. (%) |  |
| --- | --- |
| Secondary cases n=74 |  |
| **Timing of first SARS-CoV-2 detection** |  |
| At enrollment (RT-PCR) | 50 (67.6%) |
| During follow-up (RT-PCR) | 4 (5.4%) |
| At study completion (serology) | 20 (27.0%) |
| **Onset of respiratory symptoms** |  |
| At enrollment | 45 (60.8%) |
| Before enrollment | 3 (4.1%) |
| After enrollment | 13 (17.6%) |
| No symptoms | 13 (17.6%) |
| **Symptom status** |  |
| ARI | 42 (56.8%) |
| Mild symptoms | 19 (25.7%) |
| Asymptomatic | 13 (17.6%) |

Of all RT-PCR positive subjects with available serology test result upon study completion (101 index cases and 40 secondary cases), 111/141 (78.7%) had SARS-CoV-2 antibodies at time of study completion (Table 5). Seroconversion rate was highest for RT-PCR positive subjects who met the ARI case definition and decreased with fewer or no symptoms (p=0.01).

**Table 5:** Seroconversion of subjects (index cases and secondary cases) with positive RT-PCR and available serology test result at study completion by reported symptom status.

|  | No. (%) |  |  |  | P-value |
| --- | --- | --- | --- | --- | --- |
|  | Total<br>n=141 | ARI<br>n=104 | Mild symptoms<br>n=30 | Asymptomatic<br>n=7 |  |
| **Seroconversion** |  |  |  |  | 0.015 |
| Yes | 111 (78.7) | 88 (84.6) | 19 (63.3) | 4 (57.1) |  |
| No | 30 (21.3) | 16 (15.4) | 11 (36.7) | 3 (42.9) |  |

Sensitivity analysis showed a household SAR of 23.5% (20/85 households; 95%-CI: 15.3-34.2%) and a per-person SAR of 11.2% (24/215 household members; 95%-CI: 7.45-16.3%) when considering the 50 household members with a positive RT-PCR at enrollment as co-primary index cases.

## DISCUSSION

Based on the interim results of this study, we estimate that SARS-CoV-2 transmission occurs in 44.4% of households in the Netherlands and Belgium and on average 27.9% of household members get infected. The SAR was higher when index cases experienced more severe symptoms, confirming earlier observations.[3, 11-13] Two-thirds of the secondary infections among household members had already occurred at enrollment, highlighting the importance of timely case identification and isolation. Interestingly, 21.3% of subjects with RT-PCR confirmed SARS-CoV-2 infection had no detectable antibodies in DBS specimens collected 4-6 weeks post-infection, suggesting an absent, weak, or delayed humoral response in these individuals.

The household SAR found in this study is at the high end of the 4% to 45% reported so far in previous studies.[3, 4, 14-23] While this may reflect true differences in local transmission dynamics across studies, a major factor determining household SAR is the intensity of sampling protocols used. Fung et al. [16] showed in a review that the SAR estimates more than doubles for studies with a RT-PCR test frequency of >2 tests compared to one test. In this study, a dense sampling and intensive follow-up design was used, combining RT-PCR screening of all symptomatic and asymptomatic household members, repeated RT-PCR testing for new onset ARI and paired antibody testing of all subjects in the study. This allowed us to detect asymptomatic infections, and those with negative RT-PCR result. Indeed, studies that used equally or more dense sampling protocols report SARs similar to our estimates.[4, 15, 17] To quantify the effect of sampling protocols on the estimated SAR, we calculated the household and per-person SAR in our study excluding asymptomatic and RT-PCR negative subjects. This would reduce the detected SAR by 9.4% and 11.3%, which is more in line with estimates from (retrospective) household studies that are based on contact tracing investigations with symptom-based RT-PCR testing alone. We therefore conclude the SAR found in our study is a more valid estimate reflecting the true household SAR. Of note, all households currently analyzed were enrolled during a period where the more transmissible mutant strains, in particular B.1.1.7 and P.1 [24-26], did not yet circulate in the Netherlands and Belgium. It is likely that household SARs will prove to be even higher for the new variants of SARS-CoV-2.

Given the high number of co-primary cases in this study and the fact that most transmission occurred within the first five days of follow-up, we infer that transmission takes place in an early stage of infection, perhaps even before testing. It is estimated that up to 44% of transmission occurs during the pre-symptomatic period in settings with substantial household clustering.[27] Currently, households only receive information about appropriate infection control measures once a first infection is confirmed, but implementing these at symptom onset –probably together with rapid antigen self-tests for the other household members - could be potentially more effective in preventing household transmission. Especially in the beginning of the pandemic test results could be delayed since large-scale testing was not available and laboratory capacity was limited.

In line with earlier findings, household contacts of index cases with more severe respiratory symptoms are at higher risk for secondary infection.[3, 16, 19, 28] In addition, our data indicate some other characteristics that might be associated with secondary infection. The secondary infection risk was higher among household members living in a large household, among male household members and among children compared to adults. Child to parent transmission was less common than parent to child transmission. However, more data are needed for confirmatory analyses.

Our study has some limitations that merit discussion. First, the first detected household case was considered the index case, but it is possible that other household members were infected concurrently by an external source (co-primary case), or that transmission occurred in the opposite direction. This could influence analysis of SAR by index or household contacts characteristics and for co-primary cases, may overestimate the within household transmission. Alternative enrollment criteria, for instance based on household exposure rather than confirmed infection, would be needed to improve differentiation between index and secondary cases and reconstruction of transmission chains. Second, we assumed that household transmission was responsible for all infections among household contacts. The household SAR could therefore be overestimated. However, the quarantine and isolation orders should have limited community exposure. Third, we might have underestimated the SAR to some extent for study-specific reasons as 1) asymptomatic or subjects with mild symptoms were less likely to show a detectable humoral response compared to subjects with ARI.[29] This suggests that some of the asymptomatic infections during follow-up were possibly missed since only subjects with ARI were tested with RT-PCR and; 2) swabs and DBS samples were self-collected which may have reduced the sensitivity. In addition, the percentage failed tests due to insufficient sampling is probably higher compared to samples taken by a healthcare worker. However, self-sampling of midnasal swabs has been shown to minimally impact detection as compared to samples collected by healthcare professionals.[30] Similarly, DBS have proven a less invasive and valid alternative for serum.[31-33] Fourth, the study population might not be completely representative for the Dutch and Belgium population which could led to an over- or underestimation of the true SAR. The percentage households with a high educational level in the study were overrepresented compared to the average Dutch and Belgium population.[34] These households may have different living circumstances compared to people with a low social-economic status. Furthermore, the percentage of healthcare workers may be overrepresented in our study population since at the start of the SARS-CoV-2 pandemic testing was almost exclusively accessible for healthcare workers only. Healthcare workers might be more aware of possible transmission routes and appropriate infection control measures. Finally, at this stage of the data collection and sample processing, numbers were insufficient to allow detailed evaluation of individual and household characteristics associated with transmission. This will be further explored once the full dataset including 272 households becomes available.

## Conclusion

These interim results show high household SAR (44.9%) and per-person SAR (27.9%) for SARS-CoV-2 within Dutch and Belgian households. Transmission was higher from symptomatic compared to asymptomatic index cases, and most transmission had already occurred early during follow-up which underlines the importance of early SARS-CoV-2 diagnosis and isolation. In almost a quarter of RT-PCR positive cases, no antibodies were detected. Household and individual characteristics influencing transmission will be further explored as more data accumulate.

## Supporting information

See supplement for more details

## Data Availability

As this is an ongoing study the data is not yet available on a research data repository.

## ACKNOWLEDGEMENTS

We would like to thank all participants of this study. In addition, we would like to thank Marion Koopmans for providing valuable input in designing this study, Felicity Chandler and Aurora Horstink for technical support and Reina Sikkema for the substantive contribution to this manuscript.

