## Supplementary material for "Transmission of SARS-CoV-2 within households: a prospective cohort study in the Netherlands and Belgium – Interim results": See supplement for more details

**Data collection**

At study enrollment, each participant of ≥16 years received instructions to install the COVapp on a mobile device. The COVapp is a custom made application compatible with Apple and Android systems, developed by the UMCU. A unique username and password providing access to the application were sent by email. Via this application, participants could fill in several questionnaires and daily symptom diaries (self-reported; fever, coughing, sore throat, cold, dyspnea, headache, muscle pain, cold shivers, fatigue, anosmia (since July 7, 2020) and diarrhea). Parents or caregivers completed the questionnaires and symptom diaries for their children aged <16 years, or for family members without access to the app. At start of the study, per household a questionnaire was filled out containing questions regarding household characteristics and living situation. A nose-throat swab (NTS), a capillary blood sample on filter paper (dried blood spot) and a baseline questionnaire capturing demographics, comorbidities and views on prevention measures were collected from each household member irrespective of symptoms. For children aged 0-2 year, also a stool sample was collected seven days after enrollment. A daily follow-up of disease symptoms started for at least 21 days. Symptom follow-up was prolonged in case of new acute respiratory illness (ARI) episode in the household emerged. At the beginning of a new ARI episode the app generated a request for a nose-throat swab from the person with ARI disease (and an additional stool sample for children 0-2 years seven days after symptom onset) and scoring lists. At the end of the ARI episode a questionnaire about disease progression and self-reported transmission prevention behaviors popped up. Follow-up ended after 21 days or 21 days after the last date of onset of symptoms in the household. Ten days later a capillary blood sample was collected for each household member again. The mobile application automatically generated requests for swabs, blood or stools during the study. The materials for self-sampling were delivered at the beginning of the study at the home address, stored in home freezers (-15 °C) during follow-up and picked-up at the end of the study by the study team. All data entered to the app were stored in an online secured web system.

**Laboratory analysis**

At arrival on the lab blood samples and swabs were stored at -80 °C instantly until further analysis. Swabs were tested separately for the presence of SARS-CoV-2 by real-time PCR as described in detail elsewhere.[1] Briefly, 100 µl of a clinical specimen was mixed with proteinase K and 10 µl of a known amount of phocine herpesvirus (DNA) and Phocine distemper virus (RNA)—to monitor the efficient extraction of nucleic acids (NA) — and incubated for 15 min at 56°C. Subsequently, NA extraction was performed by using the specific A protocol on the NucliSens EasyMag (BioMérieux, Marcy-l'Etoile, France). The nucleic acid was then eluted in 70 µl of elution buffer and directly used for cDNA synthesis and real-time TaqMan PCR. Stool samples were mechanically pretreated by beating them through a membrane prior to the proteinase K pretreatment to optimize nucleic acid extraction. A 20-µl reaction contained 5 µl of NA extract and 5 µl of the TaqMan Fast Virus 1-Step Master Mix 4x (Thermo Fisher, Brussels, Belgium). Primer and probe sequences, as well as optimized concentrations were used as described by Corman et al.[1] All oligonucleotides were synthesized and provided by Tib Molbiol, Berlin, Germany. Thermal cycling was performed at 55°C for 10 min for reverse transcription, followed by 95°C for 3 min and then 45 cycles of 95°C for 15s, 58°C for 30s on a LightCycler 480 instrument (Roche) for the detection of the SARS-CoV-2 E-gene. In addition, NA extracts were analysed for the presence of RNAseP to check the quality of the specimens. Cycle threshold values (Ct-values) >35 were designated as invalid. The 20-µl reaction contained also 5 µl of NA extract and 5 µl of the TaqMan Fast Virus 1-Step Master Mix 4x (Thermo Fisher). Primer and probe sequences, as well as optimized concentrations were used as described by the CDC.[2] Thermal cycling was performed at 50°c for 15 min for reverse transcription, followed by 2 min at 95°C and then 45 cycles of 95°C for 15 sec and 30 sec at 55°C on the LightCycler 480 instrument (Roche). Ct-values for SARS-CoV-2 ≤40 is used as cut-off to interpreted positively.

Dried blood spot specimens were tested by multiplex protein microarray for antibodies targeting recombinant SARS-CoV-2 spike (S) ectodomain and S1 subunit antigens expressed in HEK293 cells as described earlier.[3-5] Briefly, antigens were printed on nitrocellulose coated slides and incubated for 1 hour at 37 °C with Blotto Blocker (Pierce, United States of America). Slides were washed with phosphate buffered saline (PBS) containing 0.1% Tween-20 between each step. DBS were eluted in PBS with 5% Tween-20 for one hour, diluted in Blotto Blocker with 0.1% Tween-20, and tested on the printed slides in a final 1:40 dilution for 1 hour at 37 degrees Celsius. This was followed by an incubation step with goat anti-human IgG (Fab specific) conjugated with AF647 (Jackson ImmunoResearch, United Kingdom). Slides were scanned using a Powerscanner (Tecan, Switzerland) and analyzed using ScanArray Express software (PerkinElmer, Waltham, USA). The use of DBS was compared to serum on the protein microarray platform by testing DBS specimens 1:40 and serial diluted serum specimens obtained from 8 individuals against a panel of endemic and epidemic human coronavirus recombinant antigens. The relative fluorescence units (RFU) of DBS specimens showed a good correlation and a similar level compared to 1:80 serum for various antigens. Specimens with a S1 antigen signal exceeding 13,000 relative RFU and a S ectodomain signal exceeding 2,000 RFU were considered positive.
