## Supplemental Table 1 for "Transmission of SARS-CoV-2 within households: a prospective cohort study in the Netherlands and Belgium – Interim results"

**Supplement table 1:** Sampling scheme and sample completeness based on the 117 included households (117 index cases and 265 household members).

| | Enrollment | | | ARI symptoms | | End of study |
|---|---|---|---|---|---|---|
| | DBS | NTS | Stool* | NTS | Stool* | DBS |
| Total samples requested | 382 | 382 | 15 | 78 | 9 | 382 |
| Missing result due to incomplete sampling n(%) | 13 (3.4%) | 13 (3.4%) | 3 (20%) | 8 (10.3%) | 2 (22.2%) | 10 (2.6%) |
| Missing result due to insufficient sampling n(%) | 97 (25.4%) | - | - | - | - | 61 (16.0%) |

*Stool samples are requested only for children aged 0-2 years 7 days after enrollment or ARI symptom onset.
